# Validation of a combined ELISA to detect IgG, IgA and IgM antibody responses to SARS-CoV-2 in mild or moderate non-hospitalised patients

**DOI:** 10.1101/2020.11.30.20229732

**Authors:** A M Cook, S E Faustini, L J Williams, A F Cunningham, M T Drayson, A M Shields, D Kay, L Taylor, T Plant, A Huissoon, G. Wallis, S Beck, S E Jossi, M Perez-Toledo, M L Newby, J D Allen, M Crispin, S Harding, A G Richter

**Author notes:** **Corresponding author** Dr Leigh J Williams, The Binding Site Group, 8 Calthorpe Road, Birmingham, B15 1QT, UK.

## Abstract

**Background:** Frequently SARS-CoV-2 results in mild or moderate disease with potentially lower concentrations of antibodies compared to those that are hospitalised. Here, we validated an ELISA using SARS-CoV-2 trimeric spike glycoprotein, with targeted detection of IgG, IgA and IgM (IgGAM) using serum and dried blood spots (DBS) from adults with mild or moderate disease.

**Methods:** Targeting the SARS-CoV-2 trimeric spike, a combined anti-IgG, IgA and IgM serology ELISA assay was developed using 62 PCR-confirmed non-hospitalised, mild or moderate COVID-19 samples, ≥14 days post symptom onset and 624 COVID-19 negative samples. The assay was validated using 73 PCR-confirmed non-hospitalised COVID-19 and 359 COVID-19 negative serum samples with an additional 81 DBSs, and further validated in 226 PCR-confirmed non-hospitalised COVID-19 and 426 COVID-19 negative clinical samples.

**Results:** A sensitivity and specificity of 98.6% (95% CI, 92.6–100.0), 98.3% (95% CI, 96.4–99.4), respectively, was observed following validation of the SARS-CoV-2 ELISA. No cross-reactivities with endemic coronaviruses or other human viruses were observed, and no change in results were recorded for interfering substances. The assay was stable at temperature extremes and components were stable for 15 days once opened. A matrix comparison showed DBS to correlate with serum results. Clinical validation of the assay reported a sensitivity of 94.7% (95% CI, 90.9-97.2%) and a specificity of 98.4% (95% CI, 96.6-99.3%).

**Conclusions:** The human anti-IgGAM SARS-CoV-2 ELISA provides accurate and sensitive detection of SARS-CoV-2 antibodies in non-hospitalised adults with mild or moderate disease. The use of dried blood spots makes the assay accessible to the wider community.

**Supplementary Material:** No

## Introduction

The COVID-19 pandemic resulting from severe acute respiratory syndrome coronavirus 2 (SARS-CoV-2) infection was declared an international public health emergency in early 2020 (1). Unlike previous coronavirus infection outbreaks, such as SARS-CoV and Middle East Respiratory Syndrome, the lack of rapid, overt symptomatic presentation in 80% of patients (2, 3), has meant seroconversion rather than molecular testing has become an established method to understand SARS-CoV-2 epidemiology.

Asymptomatic or even mild infection cases represent a challenge for in vitro SARS-CoV-2 diagnostic assays, both in terms of understanding the required sensitivity and specificity (4), and for obtaining samples from molecular tested positive patients. In addition, as seroconversion to different immunoglobulin types is not a synchronous process, testing of the major humoral immune system components may be preferable to limiting assessment to individual isotypes such as IgG, IgA & IgM (IgGAM).

Here we describe the validation of an ELISA, utilising recombinant SARS-CoV-2 trimeric spike glycoprotein (5, 6), with targeted detection of IgG, IgA and IgM antibodies in non-hospitalised adults with mild or moderate disease.

## Materials & Methods

### Samples

Serum and Dried Blood Spot (DBS) samples were collected from consenting non-hospitalised polymerase chain reaction (PCR) positive adults, presenting with mild or moderate symptoms of COVID-19 disease, ≥14 days post symptom onset, and from individuals COVID-19 negative (7, 8). Serum samples were stored at -20°C and thawed before analysis.

Residual anonymised samples were used in this study from several sources. For the specificity and sensitivity analysis residual samples were from commercial sources (BioIVT, approved by Western Institutional Review Board - IRB20161665; TCS Bioscience approved by The Diagnostics Investigational Review Board-IRB112015-01.1; anonymised normal samples provided by Plasma Services Group were collected with patient consent via an FDA registered blood establishment (registration number 3005275238)) and further residual samples collected in accordance with REC 2002/201; South Birmingham Research Ethics Service. For dried blood spot comparison, residual anonymised samples were collected from University of Birmingham (REC 2002/201; South Birmingham Research Ethics Service). For cross reactivity analysis, samples were provided by the Amsterdam University Medical Center of the University of Amsterdam, the Netherlands (approved by the Medical Ethics Committee; MEC 07/182MEC 07/182).Samples were used in accordance with the world medical association declaration of Helsinki Ethical principles for Medical Research involving human subjects.

## ELISA

A stabilised trimeric spike protein preparation with 95% purity (6), assessed by silver stain, was coated onto ELISA plates (Nunc High Bind, Thermo Fisher Scientific, Loughborough, UK) at 1.0 µg/ml in PBS for 24hr. Plates were blocked using 2% BSA blocking reagent (Sigma, Gillingham, UK) prepared in PBS-0.1% Tween 20 and incubated for 1hr at room temperature. Subsequently, plates were washed three times with PBS-1% Tween + 0.08% Proclin300 and air dried before packing in airtight in aluminium-coated packs with drying agent (IDS, Gateshead, UK).

High, low, negative and cut-off calibrator controls were generated using COVID-19 negative pooled normal human and specific human monoclonal IgG1, IgA and IgM anti-SARS-CoV-2 S glycoprotein (CR3022, Absolute Antibodies, Upper Heyford, UK) (9). 100 µl of either the controls (neat dilution), calibrator (neat dilution) or serum samples (diluted 1/40) were added to the coated plate and incubated for 30 minutes at room temperature. The plate was then washed three times with PBS-0.1% Tween and purified horseradish peroxidase (HRP)-labelled polyclonal rabbit antibody to human IgG, IgA, IgM (DAKO, USA) (100 µl) was added to each well and then incubated for a further 30 minutes at room temperature (RT). The plate was washed a further three times and substrate solution containing 3,3’5,5’-tetramethylbenzidine (100 µl, Surmodics Inc, Eden Prairie, MN, USA) was added to each well and then incubated at RT in the dark for 10 minutes. The reaction was stopped by the addition of phosphoric acid (100 µl) and the plate read at 450nm using an automated liquid handler (Dynex Technologies, USA).

### Pilot study: sensitivity, specificity and cut-off co-efficient

COVID-19 negative samples (n=624, median 63 years of age, range 18-98) and PCR-confirmed non-hospitalised, mild or moderate COVID-19 samples, ≥14 days post-symptom onset (n=62, median 44 years of age, range 19-63) were analysed using the human anti-IgGAM SARS-CoV-2 ELISA. The cut-off coefficient was determined, and the clinical study powered.

### Validation of the manufacturing process Sensitivity and specificity of the ELISA

Sensitivity and specificity of the human anti-IgGAM SARS-CoV-2 ELISA was assessed using COVID-19 negative serum samples (n=359), and PCR-confirmed non-hospitalised, mild or moderate COVID-19 samples (n=73), ≥14 days post symptom onset.

### Precision of the ELISA

Precision was measured in accordance with the Clinical and Laboratory Standards Institute guidelines EP12-A2 (10). Forty COVID-19 negative serum samples replicates were spiked (<1%) with specific human monoclonal IgG1 anti-SARS-CoV-2 S glycoprotein (CR3022), at C5, C50 and C95. The C50 level was defined as a response of 1.0, C5 0.8 and C95 1.2, +/-5% of the target response. Acceptance criteria was defined as, C5: ≥36 negatives out of 40 replicates; C50: 14-26 positives out of 40 replicates; C95: ≥36 positives out of 40 replicates. The coefficient of variation (% CV) was additionally calculated for each cut-off.

### Interference with the ELISA

Interference analysis was performed by spiking haemoglobin (2000 mg/L), bilirubin (200 mg/L), triglyceride (3000 mg/L) into quintuplicate serum samples that were either antibody positive or antibody negative, (normal human serum spiked with/without CR3022). Controls were generated by spiking samples with equivalent volumes of saline. The assay was deemed to have passed the interference assessment if there was no change in the reportable value following the addition of the interfering agent.

### Cross reactivity of the ELISA with other respiratory diseases

Cross-reactivity of the assay was assessed in patients with known respiratory diseases; endemic coronaviruses (n=11), influenza and parainfluenza (n=14), enteroviruses (n=5), Epstein Bar virus (n=4), adenovirus (n=1) or respiratory syncytial virus (n=4). Positive cross-reactivity was identified if the cut-off index value was ≥1.

### Stability of the ELISA

Extremes of temperature stability was determined in triplicate following 24 hour storage at either 37°C or -20°C. Open vial stability was assessed over 15 days with 5 day interval sampling. Briefly, ELISAs were opened in parallel and components re-capped and stored at 4°C.

### Dried blood spot versus serum samples

Paired DBS and serum samples (49 PCR-confirmed non-hospitalised, mild or moderate COVID-19 samples and 32 COVID-19 negative samples), were assayed using the human anti-IgGAM SARS-CoV-2 ELISA. Capillary blood samples (50 µl per spot) were collected onto 226 grade forensic collection cards (Ahlstrom Munksjo, Germany). Individual 12 mm diameter pre-perforated DBS spots were isolated using a sterile pipette tip and placed into a universal tube at a ratio of one spot to 250 µL 0.05% PBS Tween-20 (Sigma). Tubes were briefly vortexed and incubated overnight at room temperature. DBS eluate was subsequently harvested into a microtube and centrifuged at 10,600 x g for 10 minutes at room temperature and stored at 4°C until used (11). DBS samples were applied to the assay at a 1 in 4 dilution, resulting in a 1 in 40 overall dilution. Results were analysed using Bland Altman, Passing-Bablok and Cohen’s kappa statistic.

### Clinical study

COVID-19 negative (n=426) and PCR-confirmed non-hospitalised, mild or moderate COVID-19 samples (n=226), ≥ 14 days post symptom onset, were assayed using the human anti-IgGAM SARS-CoV-2 ELISA.

### Statistics

All graphs and statistical analysis were generated using Graph Pad Prism statistical software version 5.04 or Analyse-it® for Microsoft Excel. Assay validation sensitivity, specificity and precision (95% CI and % CVs) were calculated using Analyse-it. The matrix comparison was analysed using Bland-Altman, Passing Bablok and Cohen’s kappa statistics (Analyse-it). To achieve an expectant 98%, minimum 93%, sensitivity in the clinical study, a 90% power with 5% alpha error was determined from the pilot study. A p<0.05 was considered statistically significant.

## Results

### Pilot study to determine sensitivity, specificity and cut-off co-efficient of the ELISA

The pilot study with 624 COVID-19 negative and 62 PCR-confirmed non-hospitalised, mild or moderate COVID-19 samples, which were at least 14 days from symptom onset were analysed on the human anti-IgGAM SARS-CoV-2 ELISA. The ELISA output result was reported as a ratio to the cut-off calibrator (run in parallel) and multiplied by the cut-off co-efficient, which was also determined from the pilot study and would maintain batch-to-batch consistency, defined here as 1.15. A sensitivity of 98.5 % (95% CI 91.8-100%) and specificity of 97.6% (95% CI 96.1-98.7%) was achieved, (Table 1 & Figure 1).

**Table 1.** Sensitivity and specificity of the pilot, validation and clinical study using the human anti-IgGAM SARS-CoV-2 ELISA. Assay assessed using COVID-19 negative samples and PCR-confirmed non-hospitalised, mild or moderate COVID-19 samples, ≥14 days post symptom onset.

| Pilot, Validation & Clinical Studies: Specificity & Sensitivity |  |  |  |  |  |  |
| --- | --- | --- | --- | --- | --- | --- |
| Study | Category | Number | Positive | Negative | Specificity (95% CI) | Sensitivity (95% CI) |
| Pilot Study | COVID-19 negative | 624 | 15 | 609 | 97.6<br>(96.1-98.5) | 98.5%<br>(91.4-99.7) |
|  | PCR confirmed, non-hospitalised COVID-19 positive | 62 | 61 | 1 |  |  |
| Validation Study | COVID-19 negative | 359 | 6 | 353 | 98.3<br>(96.4-99.4) | 98.6<br>(92.6-100.0) |
|  | PCR confirmed, non-hospitalised COVID-19 positive | 73 | 72 | 1 |  |  |
| Clinical Study | COVID-19 negative | 426 | 7 | 419 | 98.4<br>(96.6-99.3) | 94.7<br>(90.9-97.2) |
|  | PCR confirmed, non-hospitalised COVID-19 positive | 226 | 214 | 12 |  |  |

**Figure 1.**
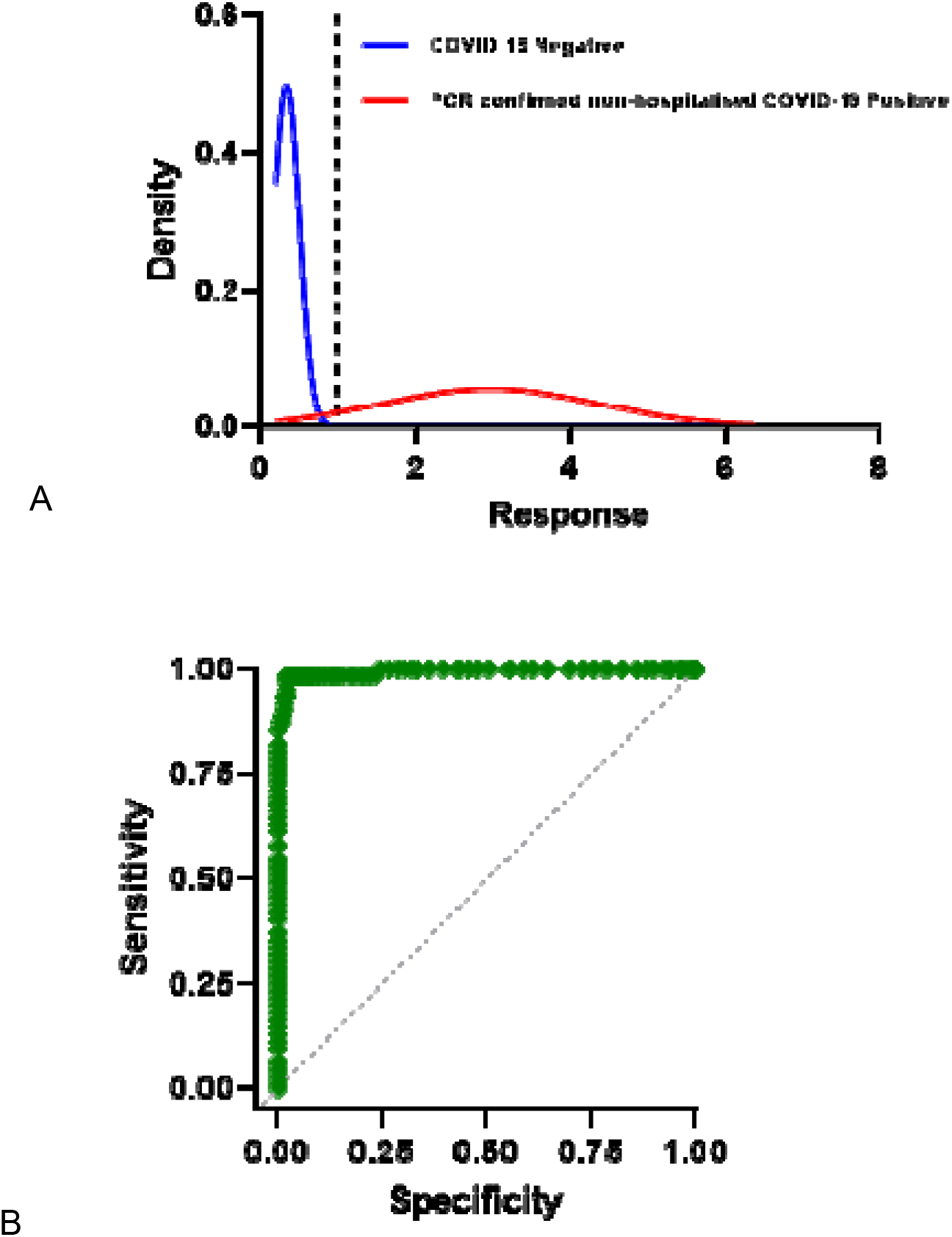
The distribution of optical densities and sensitivity and specificity of the human anti-IgGAM SARS-CoV-2 ELISA. The cumulative standard normal distribution of optical densities of (A) COVID-19 negative samples (n=624, blue) and PCR-confirmed non-hospitalised, mild or moderate COVID-19 samples, ≥14 days post symptom onset (n=62, red). (B) ROC curve of sensitivity (98.4%, 95% CI 91.4-99.7%) and specificity (97.6%, 95% CI 96.1-98.5%).

From the pilot study a minimum of 167 PCR-confirmed non-hospitalised, mild or moderate COVID-19 samples, ≥14 days post symptom onset would be required for the clinical study to determine whether the ELISA test delivered the expected sensitivity of 98% (minimum of 93%) with an alpha error of 5% and power of 90%.

### Validation of the manufacturing process

Sensitivity and specificity were assessed in COVID-19 negative samples (n=359) and PCR-confirmed non-hospitalised, mild or moderate COVID-19 samples, ≥14 days post symptom onset (n=73). COVID-19 negative samples were identified as negative for SARS-CoV-2 antibodies when assay results were less than 1 and PCR confirmed COVID-19 positive samples were identified as positive when the assay results were greater than 1, using the human anti-IgGAM SARS-CoV-2 ELISA (Figure 2). This resulted in an assay sensitivity of 98.6% (95% CI 92.6-100.0%) and specificity of 98.3% (95% CI 96.4-99.4%), (Table 1).

**Figure 2.**
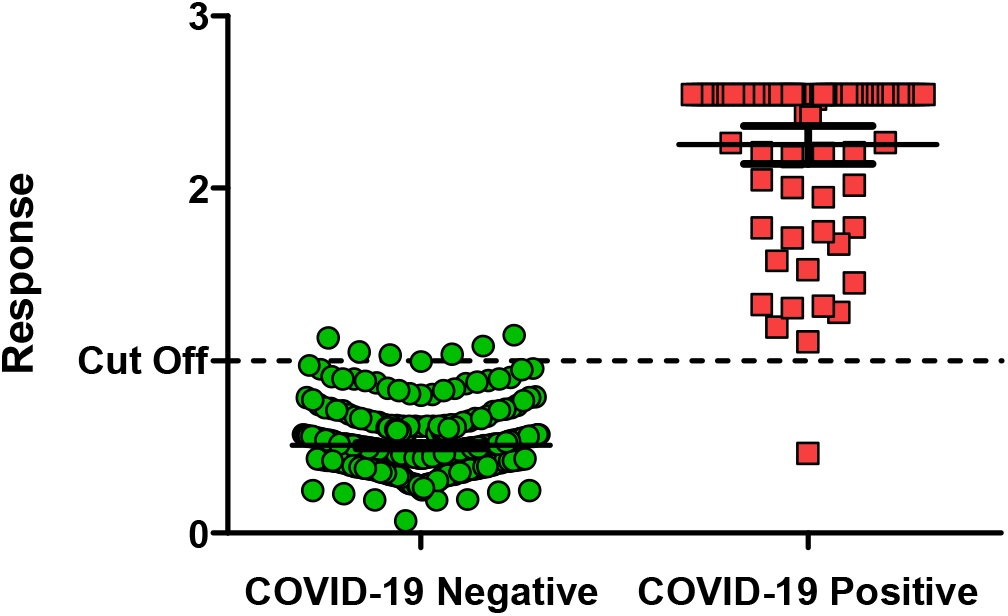
IgGAM response in COVID-19 negative (n=359) and PCR-confirmed non-hospitalised, mild or moderate COVID-19 samples, ≥14 days post symptom onset (n=73). A sensitivity and specificity of 98.6% (95% CI 92.6-100%) and 98.3% (95% CI 96.4-99.4%), respectively, was reported using an assay cut-off of 1.0.

Assay precision was assessed in 40 replicates by following the CLSI EP12-A2 guidelines to determine whether the C5 to C95 response range (range of responses at which imprecision around the C50 occurs) falls within 20% of C50. The precision of the assay was demonstrated at +/-20% of C50 as C5 gave a mean of 0.77 (40 negatives out of 40 replicates, 4.56% CV), C50 gave a mean of 1.02 (25 positives out of 40 replicates, 5.07% CV) and C95 gave a mean of 1.13 (40 positives out of 40 replicates, 3.97% CV), (Figure 3).

**Figure 3.**
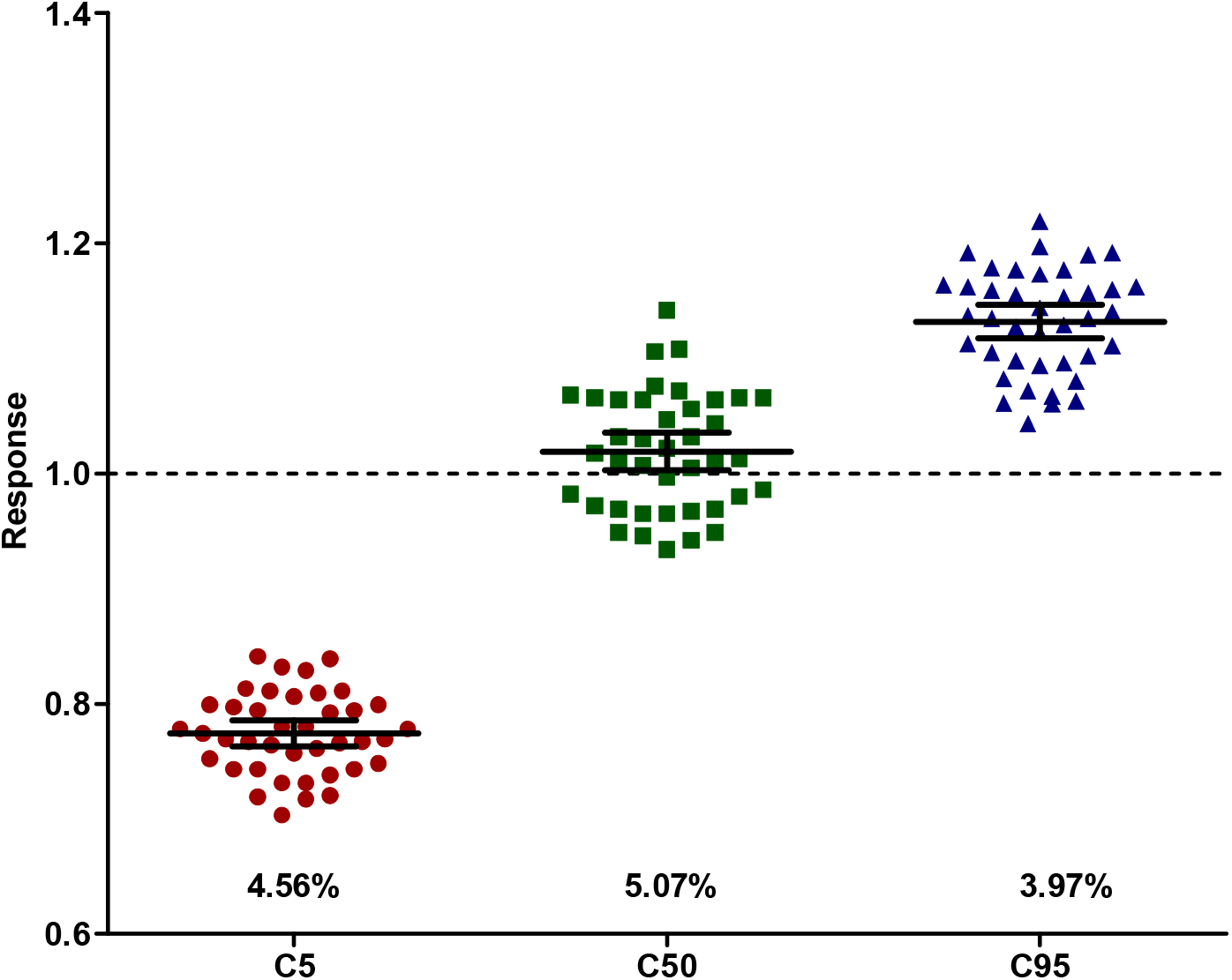
Precision of the human anti-IgGAM SARS CoV-2 ELISA. Precision was measured using clinical and lab standards guidelines EP12-A2 in 40 replicates and assessed at clinical cut-offs C5, C50 and C95. The mean and coefficient of variation (%) was calculated for each clinical cut-off.

No change in result outputs were observed following potential analytical interference from increased haemoglobin, bilirubin or triglyceride concentrations, (Figure 4 A). Assessing cross-reactivity with other human viruses and coronaviruses, 39 samples tested negative covering 8 disease categories, (Figure 4 B). Kit components were stable once opened for 15 days and for one day at extremes of temperature (Figure 4 C & D).

**Figure 4.**
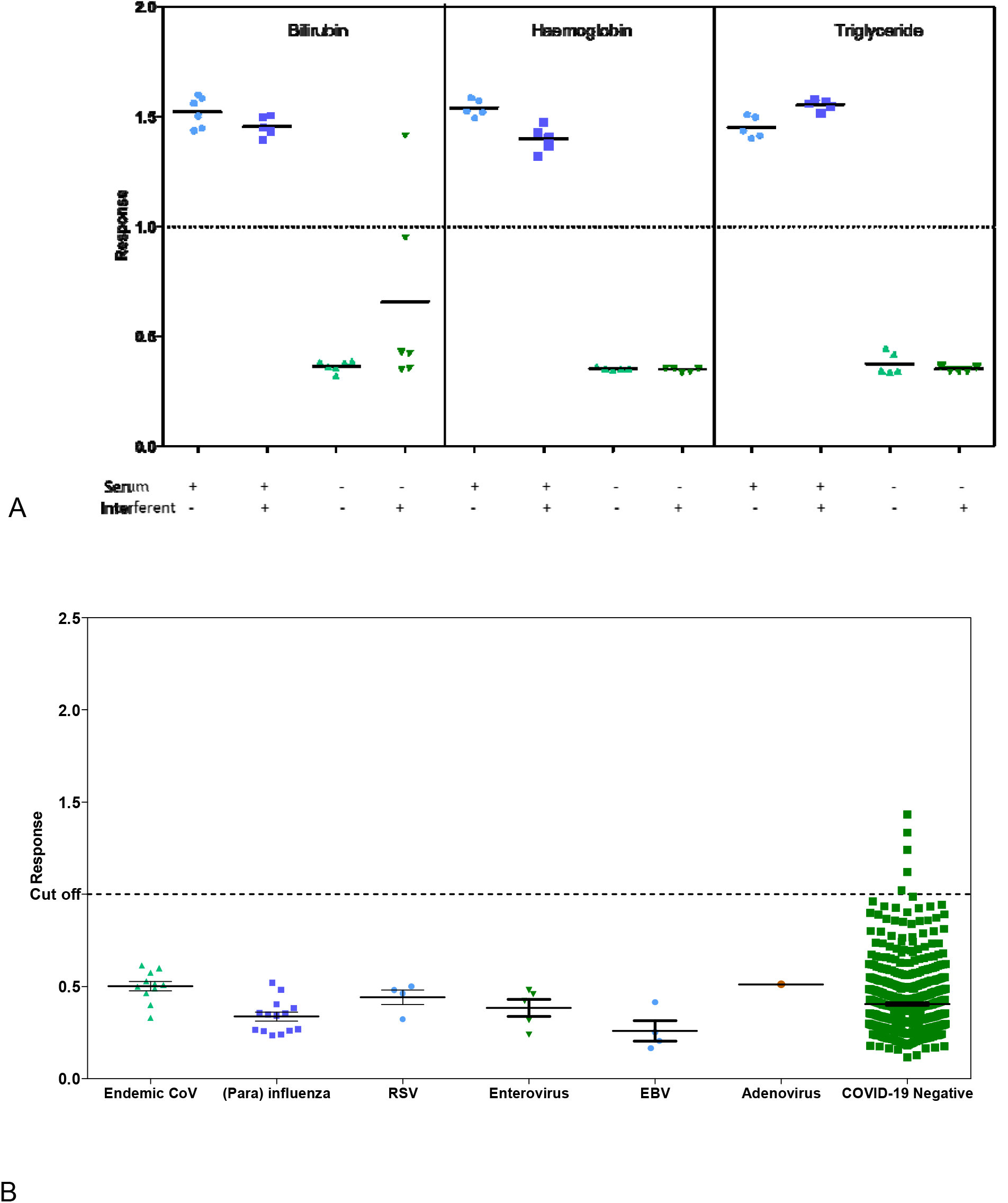
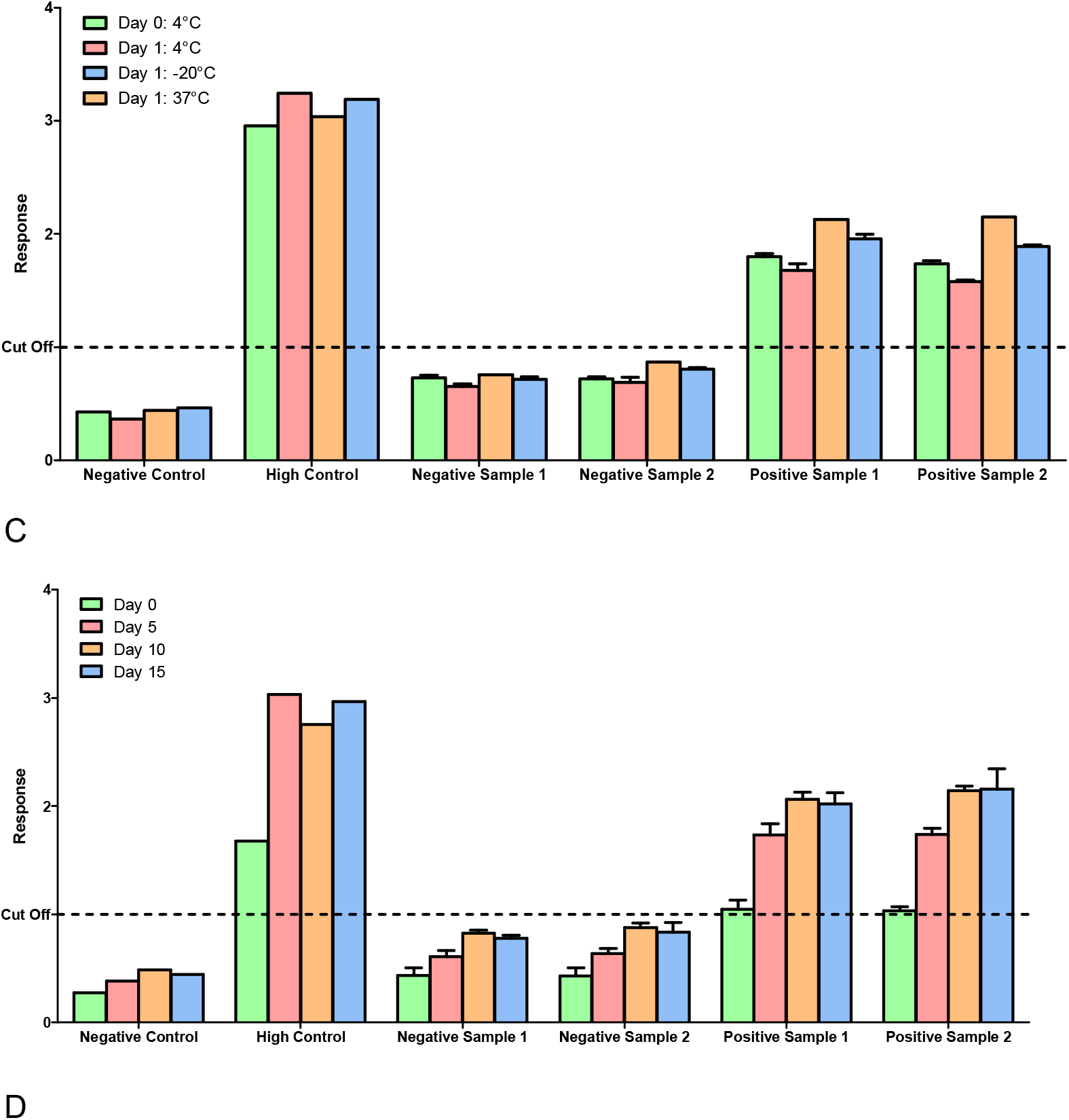
Stability and cross reactivity of antibody positive or antibody negative serum samples on the anti-IgGAM SARS CoV-2 ELISA. The assay was assessed for (A) interfering substances, bilirubin, haemoglobin and triglyceride (n=5 replicates per test) in antibody positive or antibody negative serum samples; and (B) evaluated for cross reactivity with human viruses and other endemic coronaviruses (n=39). Additionally, the kit components were tested for (C) stability at extreme temperatures (−20°C, 4°C and 37°C) (n=3) and (D) stability once opened (n=4).

### Dried blood spot versus serum samples

Responses were measured in 81 matched serum and DBS samples. A strong correlation was observed between the two matrices when assessed using the ELISA (Passing Bablok r=0.959, y=0.16-0.91x), with a mean positive bias towards the DBS compared to serum (Bland Altman 0.05, 95% CI -0.773 to 0.679), (Figure 5). An almost perfect agreement, 0.83 (Cohen’s kappa) was observed between the two sampling matrices.

**Figure 5.**
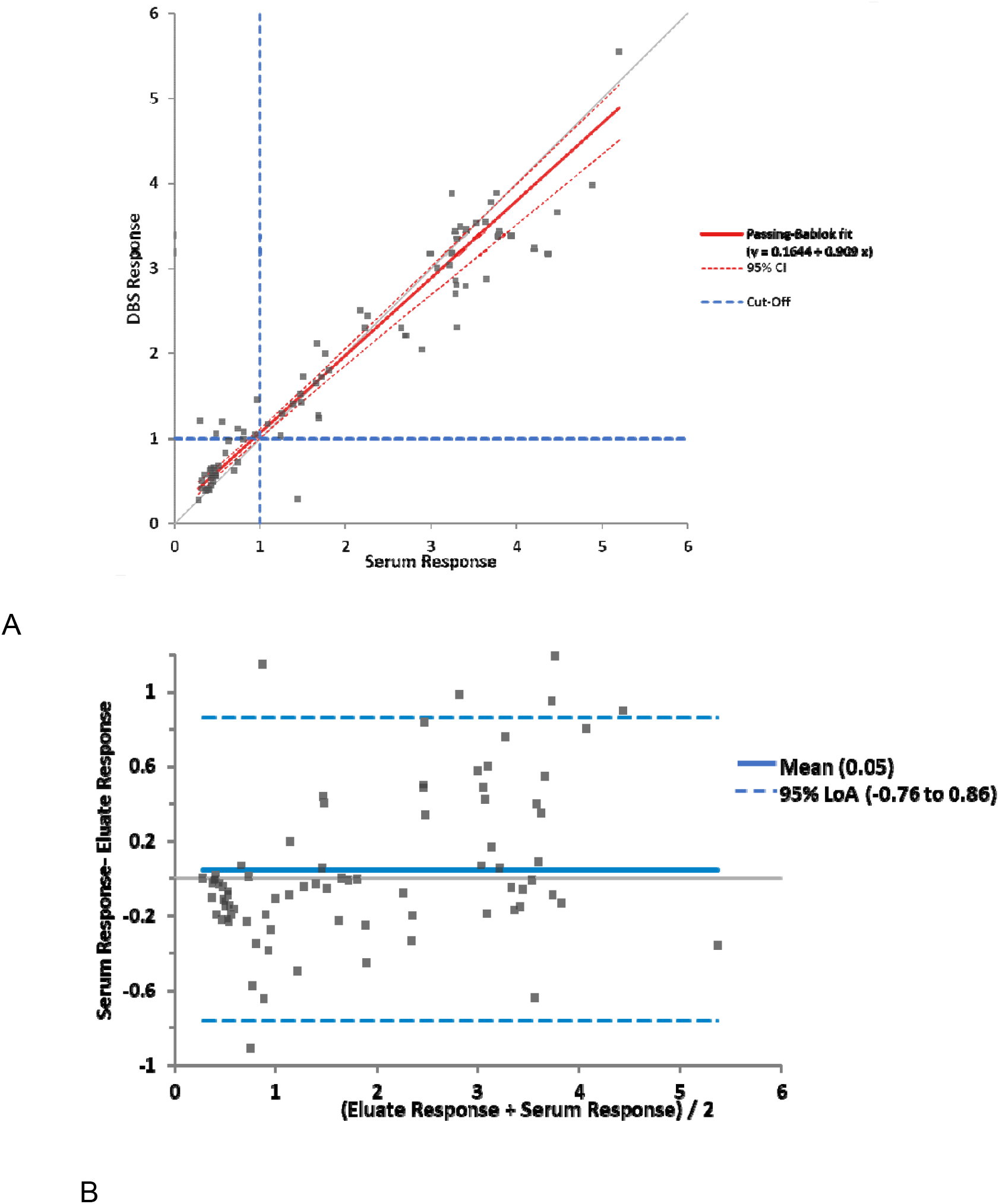
Comparison of the anti-IgGAM SARS-CoV-2 ELISA response in DBS and serum samples. The response was measured in 81 DBS and serum matched samples. A comparison between the two collection methods were assessed using (A) Passing-Bablok analysis (r=0.959, y=0.16+0.91x) and (B) Bland-Altman analysis (central line demonstrates mean difference and broken lines shows the limits of agreement, -0.76 to 0.86).

### Clinical study

The assay was validated in clinical samples from an independent group of COVID-19 negative samples (n=426) and PCR-confirmed non-hospitalised, mild or moderate COVID-19 samples, ≥14 days post symptom onset (n=226). The sensitivity of the anti-IgGAM SARS-CoV-2 ELISA in this cohort was 94.7% (95% CI, 90.9-97.2%) and specificity 98.4% (95% CI, 96.6-99.3%), (Figure 6 & Table 1).

**Figure 6.**
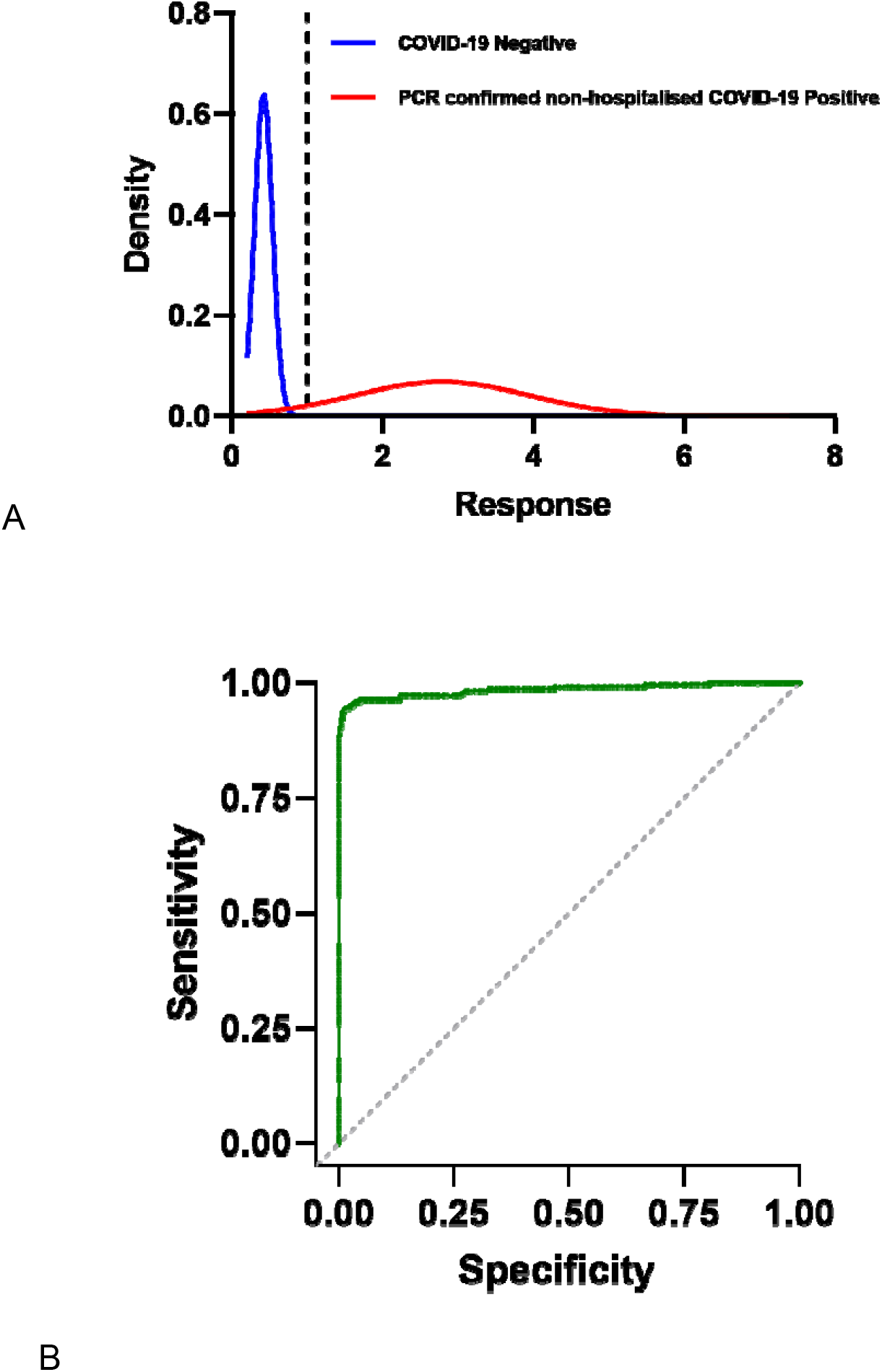
Clinical study. The cumulative standard normal distribution of (A) COVID-19 negative samples (n=426, blue) and PCR confirmed, mild or moderate COVID-19 positive samples,≥14 days post symptom onset samples (n=226, red). (B) ROC curve of sensitivity (94.7%, 95% CI 90.9-97.2%) and specificity (98.4%, 95% CI 96.6-99.3%).

## Discussion

Seroepidemiological studies are essential in understanding population-level exposure to SARS-CoV-2, determining correlates of protection from re-infection and guiding public health policy. The accuracy of such studies is determined by the performance characteristics of the immunoassay used to determine serostatus. Using samples derived from non-hospitalised adults with mild or moderate COVID-19 disease we developed a highly sensitive (98.6%) and specific (98.3%) anti-IgGAM SARS-CoV-2 ELISA. The assay showed no cross reactivity with endemic coronaviruses or other respiratory diseases, even though 30% of common colds are caused by alpha and beta coronaviruses (12)

By validating this assay using samples from non-hospitalised adults, we demonstrate the human anti-IgGAM SARS-CoV-2 ELISA to be a reliable assay for future seroepidemiological studies of the general population, the majority of whom will not require hospitalisation following infection with SARS-CoV-2. Current testing in the community has been problematic due to the inadequate sensitivities and specificities of current point of care tests in non-hospitalised patients with mild or moderate disease (4).

The assay targets nearly the entirety of the trimeric spike protein, and thereby offers a greater range of epitopes, as reports suggest neutralising antibodies to SARS-CoV-2 are not only targeted towards the receptor binding domain but also elicit high affinity and cross-reactivity to other sites across the spike (5, 13-16). Faustini et al. compared the receptor binding domain, S1 domain and nucleocapsid and although these targets were excellent at detecting antibodies in severe COVID-19 patients, they were not as effective when antibody concentrations were more limited (17). Serological assays have been developed to detect the antibody response present in severe disease (hospitalised patients). This assay was therefore developed and tested in non-hospitalised adults, at least 14 days post-symptom onset, with mild or moderate COVID-19, where the antibody concentrations are much lower than in hospitalised patients with severe disease (4, 17, 18).

The assay measures anti-IgG, IgA and IgM as reports suggest an asynchronous serological response. In adults, Seow et al. showed that the kinetics of IgGAM only occurred synchronously in 50% patients, with 9.7% developing either IgG, A and M first, 9.7% developing IgM and IgG together, 3% developing IgA and IgM together, and 3% developing IgA and IgG together (19). Valdivia et al. reported that 3.5% asymptomatic patients presented with IgM while negative for IgG (20). Long et al. showed that seroconversion for IgG and IgM occurred synchronously or simultaneously (21). Additionally, the longevity of the antibody responses is still under investigation. Studies by Iyer et al. showed that adult patients were IgA and IgM seronegative by 51 and 47 days, respectively, while IgG persisted to the end of their evaluation period (75 days) (22). Gudbjartsson, reported IgG persisting without reduction up to 4 months after diagnosis (23).

This assay benefits by being able to assess SARS-CoV-2 IgGAM antibodies in both mild and moderate disease, while also being compatible with remote sampling. Here, antibodies obtained from DBSs were shown to correlate well with serum results. DBSs are readily used in analytical assays for viruses such as those for human immunodeficiency virus (HIV) and hepatitis B and C (24-26) They are also frequently used for other applications such as screening of neonates, metabolic profiling, pharmacokinetic, forensic and environmental assays (24, 27, 28). The major advantages with using DBS’s include: the requirement for a smaller volume of blood, reduced risk of potential bacterial contamination, non-invasive blood collection, easy delivery-to-test centre, low cost and stability for prolonged periods of time with minimal deterioration of analytes (11, 24, 27). The additional advantage in the current COVID-19 pandemic include, ensuring patients do not have to attend hospitals and sites with greater risk of infection. Rickman et al. reported in a major London Teaching Hospital 15% of in-patient COVID-19 cases developed as a result of varied transmission routes within the hospital (29) and Meredith et al. also reported strong and plausible epidemiological links following the identification of 35 clusters of identical viruses associated amongst patients in their hospital (30). Therefore, when using DBSs, the test becomes accessible to all, including those that are vulnerable and those beyond developed countries.

In conclusion, utilising the trimeric spike protein and measuring IgG, IgA and IgM simultaneously, the human anti-IgGAM SARS-CoV-2 ELISA assay provides accurate and sensitive detection of antibody responses to SARS-CoV-2 in adults with mild or moderate disease. The use of dried blood spots makes this assay more accessible to the wider community.

## Data Availability

Due to the nature of this research, participants of this study did not agree for their data to be shared publicly, so supporting data is not available.

## Declarations

LJW, AMC, GW, SH & DK are members of The Binding Site Group Ltd.

## Acknowledgments

We thank Lia van der Hoek for seasonal coronaviruses samples used in the cross reactivity study from the Amsterdam Cohort Studies (ACS) on HIV infection and AIDS. The ACS is a collaboration among the Public Health Service of Amsterdam, the Amsterdam University Medical Center of the University of Amsterdam, the Sanquin Blood Supply Foundation, the Medical Center Jan van Goyen and the HIV Focus Center of the DC-Clinics. It is part of the Netherlands HIV Monitoring Foundation and financially supported by the Center for Infectious Disease Control of the Netherlands National Institute for Public Health and the Environment. The authors thank all ACS participants for their contribution, as well as the ACS study nurses, data managers and lab technicians.

## List of Abbreviations

DBS: Dried Blood Spots
ELISA: Enzyme Linked Immunosorbent Assay
PCR: Polymerase Chain Reaction
SARS-CoV-2: Severe Acute Respiratory Syndrome Coronavirus 2

